# Intraoperative microseizure detection using a high-density micro-electrocorticography electrode array

**DOI:** 10.1101/2021.09.13.21263449

**Authors:** James Sun, Katrina Barth, Shaoyu Qiao, Chia-Han Chiang, Charles Wang, Shervin Rahimpour, Michael Trumpis, Suseendrakumar Duraivel, Agrita Dubey, Katie E. Wingel, Iakov Rachinskiy, Alex S. Voinas, Breonna Ferrentino, Derek G. Southwell, Michael M. Haglund, Allan H. Friedman, Shivanand P. Lad, Werner K. Doyle, Florian Solzbacher, Gregory Cogan, Saurabh R. Sinha, Sasha Devore, Orrin Devinsky, Daniel Friedman, Bijan Pesaran, Jonathan Viventi

## Abstract

One-third of epilepsy patients suffer from medication-resistant seizures. While surgery to remove epileptogenic tissue helps some patients, 30–70% of patients continue to experience seizures following resection. Surgical outcomes may be improved with more accurate localization of epileptogenic tissue. We have previously developed novel thin-film, subdural electrode arrays with hundreds of microelectrodes over a 100–1,000 mm^2^ area to enable high-resolution mapping of neural activity. Here we used these high-density arrays to study microscale properties of human epileptiform activity. We performed intraoperative micro-electrocorticographic recordings within epileptic cortex (the site of seizure onset and early spread) in nine patients with epilepsy. In two of these patients, we obtained recordings from cortical areas distal to the epileptic cortex. Additionally, we recorded from two non-epileptic patients with movement disorders undergoing deep brain stimulator implantation as non-epileptic tissue controls. A board-certified epileptologist identified microseizures, which resembled electrographic seizures normally observed with clinical macroelectrodes. Epileptic cortex exhibited a significantly higher microseizure rate (2.01 events/min) than non-epileptic cortex (0.01 events/min; permutation test, *P*=0.0068). Using spatial averaging to simulate recordings from larger electrode contacts, we found that the number of detected microseizures decreased rapidly with increasing contact diameter and decreasing contact density. In cases in which microseizures were spatially distributed across multiple channels, the approximate onset region was identified. Our results suggest that micro-electrocorticographic electrode arrays with a high density of contacts and large coverage are essential for capturing microseizures in epilepsy patients and may be beneficial for localizing epileptogenic tissue to plan surgery or target brain stimulation.

## Introduction

Epilepsy affects 1% of the global population, and drugs alone fail to control seizures in ∼30% of cases.^1,2^ Despite the continued development and approval of novel anti-seizure medications, the high prevalence of drug-resistant epilepsy has persisted for several decades.^2-4^ Some patients with drug-resistant, focal-onset epilepsy benefit from surgical resection or ablation of epileptogenic tissue.^1,5-7^ Although this treatment is a valuable alternative, surgical resection yields complete postoperative seizure freedom in 30–70% of patients, depending on factors such as a patient’s neuropathology, the presence, type, and localization of structural lesions, and the timing and extent of resection.^4,6^ Resective and ablative surgeries also carry risks of post-operative neurocognitive deficits that are in part related to the amount of tissue resected.^4,8^ Furthermore, focal epilepsy patients are not candidates for surgical resection or ablation if they exhibit wide-spread or multifocal epilepsy networks, overlap of epileptogenic cortex with eloquent brain areas, or serious medical comorbidities.^4^ Neuromodulation therapies such as responsive neurostimulation (RNS), deep brain stimulation (DBS), and vagus nerve stimulation (VNS) offer alternatives for these patients but seldom produce lasting seizure freedom.^4,9,10^

Resection, ablation (e.g., laser interstitial thermal or radio-frequency ablation), and RNS require precise targeting of the epileptogenic zone (EZ)—the brain region for which removal is necessary and sufficient to control seizures.^1,4,11-14^ Diagnostic tools currently used to identify the EZ include neuroimaging, seizure semiology, scalp electroencephalography (EEG), electrocorticography (ECoG), and stereo-electroencephalography (sEEG).^1,12,15^ However, these tools suffer from a lack of spatial precision.^16-19^ Invasive neurophysiological techniques such as ECoG and sEEG record aggregate activity from ∼5 mm^2^ of tissue, including local neuronal firing, intrinsic currents, and synaptic potentials from near and distant sources.^20,21^ Spatially-localized epileptiform activity on the submillimeter scale is not apparent in standard clinical recordings, but has been revealed by experimental recordings from epileptic brain tissue using silicon shank microelectrode arrays and surface and intraparenchymal microwire electrode arrays.^16,22^ Recordings in epileptic patients using microwire electrode arrays have identified seizure-like discharges isolated to a single 40-µm wire contact, termed “microseizures”.^16^ These microseizure discharges sometimes evolve into clinical seizures involving multiple square centimeters of cortex.^16^ While the presence of spontaneous microseizures indicates that ictal activity may begin at the microscale, the relevance of these events to the organization of the EZ and to clinical seizure onset remains unknown.

One limitation of microarrays that have previously been used to detect microseizures is their minimal coverage of the cortical surface. Here we leveraged advances in micro-electrocorticographic (µECoG) electrode arrays with broad, high-density microcontact coverage to study the microscale dynamics of epileptiform activity in human epilepsy patients.^23^ Our µECoG recordings targeted areas of cortex with clinically identified seizure onset or early spread—an approximation of the EZ which we refer to as epileptic cortex (EC). For controls, we performed µECoG electrode array recordings outside of the EC as well as in movement disorder patients undergoing DBS implantation. We hypothesized that microseizures occur more frequently in EC than in non-epileptic cortex, and that most microseizures are spatially restricted to <1–2 mm^2^, thus precluding their detection by standard clinical recording electrodes. Our µECoG electrode arrays were minimally invasive and consisted of a flexible, liquid crystal polymer, thin-film (LCP-TF) substrate with electroplated gold (Au) or platinum-iridium (PtIr) microcontacts.^23^ The scale and configuration of the LCP-TF µECoG electrode arrays offered high-density microscale recordings (0.762–1.72 mm spacing) while maintaining adequate spatial coverage of the cortical surface (144–798 mm^2^), an important compromise between the two spatial extremes of clinical ECoG arrays (10-mm spacing, ≤6,400-mm^2^ coverage) and silicon shank microelectrode arrays (0.4-mm spacing, 16-mm^2^ coverage).^21,22^ The LCP-TF fabrication method created a smooth array surface which minimized the possibility of tissue damage due to surface penetration, which occurs when using silicon shank or microwire electrodes.^23^ Our objectives were to determine whether microseizures can be readily observed in brief intraoperative recordings, to assess the specificity of microseizures for EC, to characterize the spatial scale of detected microseizures, and to determine whether microelectrode recording devices are necessary for their detection. Our results indicate that microseizures observed using LCP-TF µECoG electrode arrays may provide a more precise tool to improve pre-surgical evaluation for drug-resistant epilepsy.

## Materials and Methods

### Participants

We performed intraoperative recordings in nine patients with drug-resistant epilepsy undergoing surgical resection or electrode removal after the EC had been clinically identified. As a control, we also recorded outside of the EC in two of the nine epileptic patients and recorded intraoperatively from the cortex of two patients with movement disorders during implantation of a DBS device. Informed consent was obtained from all patients in accordance with and ethical approval was granted by the Institutional Review Boards at New York University Langone Health and the Duke University Health System.

### LCP µECoG arrays

LCP µECoG arrays were fabricated by Dyconex (Bassersdorf, Switzerland). The arrays have passed ISO 10993 biocompatibility tests for hemolysis, cytotoxicity, and material-mediated pyrogenicity at a contract research organization for medical devices (NAMSA; Toledo, USA) following FDA Good Laboratory Practice regulations. We used LCP-TF arrays with three different designs: 128 microcontacts with 1.33-mm center-to-center spacing (pitch), 244 microcontacts with 0.76-mm pitch, and 256 microcontacts with 1.72-mm pitch (Fig. 1A).^23^ All microelectrode contacts were 200 µm in diameter. The back of the LCP-TF arrays were coated in polydimethylsiloxane (Dow Corning MDX4-4210, USP Class VI) to allow for mechanical configuration of the arrays and to prevent possible tissue damage by the thin edges of the LCP-TF (Fig. 1C).^23,24^ The 128-contact design was sufficiently narrow to permit subdural implantation via a burr-hole during DBS surgery for movement disorders. Electrodes were connected to custom modular Intan headstages for amplification and digital sampling at 20 kilo samples per second (kSPS) (Fig. 1B). The array, headstages, and cabling were sterilized with ethylene oxide gas at Duke or with hydrogen peroxide (STERRAD) at NYU.

**Figure 1.**
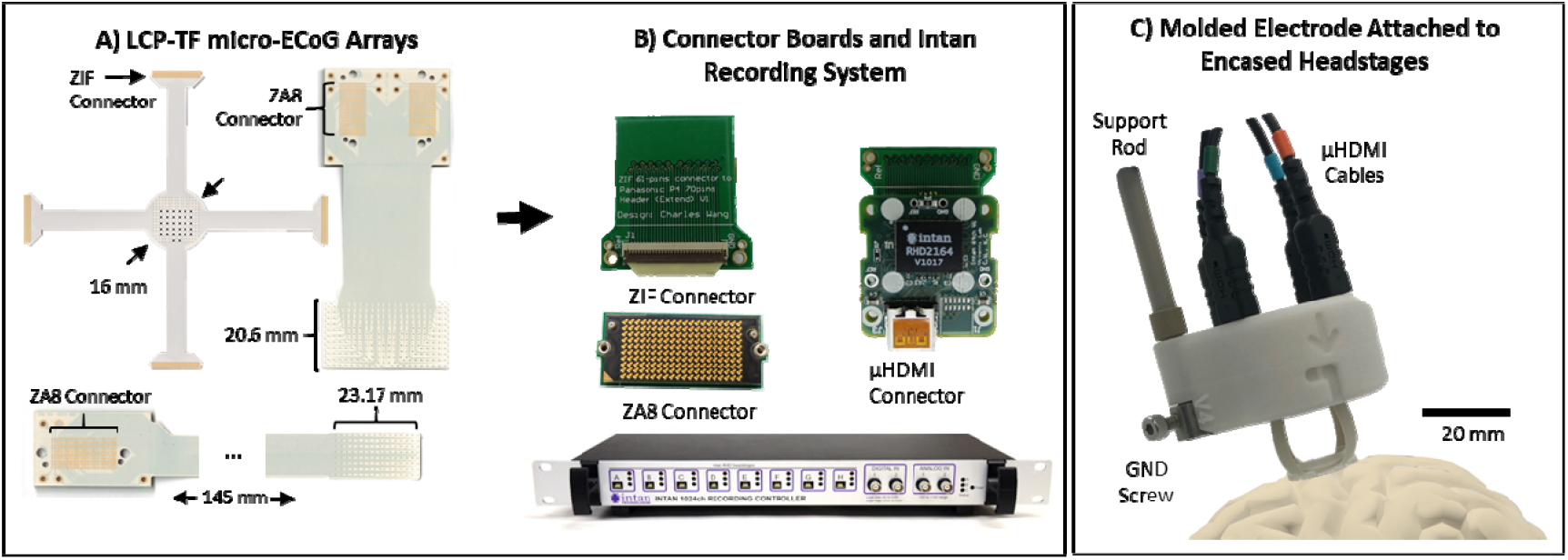
LCP-TF electrodes and intraoperative recording system. A) Flexible liquid crystal polymer (LCP) electrodes with 244 (left), 256 (right), and 128 (bottom) Au recording contacts (200-µm diameter; pitch = 0.762 mm [left], 1.72 mm [right], 1.33 mm [bottom]). B) Custom digitizing headstage using an Intan Technologies integrated circuit. Electrode arrays were connected to the digitizing headstage using either Zero Insertion Force (ZIF) or Samtec ZA8 adaptor printed circuit boards. The Intan Technologies recording controller collected digital signals from the headstages. C) Example of an LCP-TF electrode array molded in silicone and attached to four headstages inside a 3D-printed support structure with µHDMI cables for connection to the recording controller. Schematic depicts electrode placement on the cortex.

### Intraoperative recordings

Recording procedures in the operating room were developed with input from neurosurgeons and neurologists. Craniotomies, burr hole drilling, and all other surgical procedures were performed solely for standard of care clinical purposes. During intraoperative recordings from epilepsy patients, the neurosurgeon placed a sterilized LCP-TF surface array within the EC, defined as the area with seizure onset or early spread as identified by a neurologist using clinical macroelectrode recordings (Fig. 1C). In two patients, a second recording was performed at a site outside of the EC. During intraoperative recordings from DBS patients, the neurosurgeon slid a 128-contact array through a burr hole along the cortical surface. Sterile alligator clips connected the ground and reference terminals of the headstages to an accessible metal connection on the patient’s body such as a metal scalp retractor or bone screw. Headstages were connected by micro high-definition multimedia interface (µHDMI) cables to an Intan Technologies recording controller that was positioned outside the sterile zone. Recordings were conducted either under anesthesia or when patients were awake during intraoperative mapping of eloquent cortex or motor control (Table 1).

**Table 1.** Clinical summary of patients.

| Patient | Age | Sex | Diagnosis | Array location | Recording duration | Channels | Anesthesia |
| --- | --- | --- | --- | --- | --- | --- | --- |
| 1 | 41-45 | F | Focal epilepsy | L posterior superior temporal gyrus | 18m 10s | 256 | Isoflurane (0.8%) |
| 2 | 31-35 | M | Focal epilepsy | L posterior superior temporal gyrus | 18m | 244 |  |
| 3 | 36-40 | M | Focal epilepsy | L posterior superior temporal gyrus | 8m 50s | 256 |  |
| 4 | 36-40 | M | Focal epilepsy | Left anterior middle temporal gyrus | 6m | 244 | Propofol (50 mcg/kg/min)<br>Remifentanyl (0.1 mcg/kg/min)<br>Dexmedetomidine (0.2 mcg/kg/hr) |
| 5 | 41-45 | F | Focal epilepsy | L supramarginal gyrus | EC: 5m 28s<br>Non-EC: 5m | 244 | Sevoflurane (1.64%)<br>Remifentanyl (0.05 mcg/kg/min)<br>Dexmedetomidine (0.2 mcg/kg/hr) |
| 6 | 46-40 | F | Focal epilepsy | L anterior inferior temporal gyrus | 10m 40s | 244 | Propofol (130 mcg/kg/min)<br>Remifentanyl (0.125 mcg/kg/min) |
| 7 | 26-30 | M | Focal epilepsy | R posterior middle temporal gyrus | EC: 5m<br>Non-EC: 5m | 244 | Propofol (100 mcg/kg/min)<br>Remifentanyl (0.1 mcg/kg/min) |
| 8 | 31-35 | M | Focal epilepsy | L middle temporal gyrus | 6m | 244 | Propofol (25 mcg/kg/min)<br>Remifentanyl (0.03 mcg/kg/min)<br>Dexmedetomidine (0.3 mcg/kg/hr) |
| 9 | 46-50 | M | Focal epilepsy | L posterior superior temporal gyrus | 16m 30s | 2 x 256 |  |
| 10 | 61-65 | F | Movement disorder | L motor cortex | 53m 40s | 128 |  |
| 11 | 71-75 | F | Movement disorder | L motor cortex | 7m 35s | 128 |  |

### Data preprocessing

Neural data was recorded at 20 kSPS, low-pass filtered using a multitaper filter with a time window of 0.01 s and frequency bandwidth of 300 Hz, and downsampled to 1 kHz. Channels with high impedance (>250 kΩ at 1 kHz on post-operative testing) and epochs with excessive artifact (based on visual inspection) were excluded from analysis.

### Data analysis

A line length (LL) detector was used to screen recordings for candidate events.^25^ LL was calculated over an interval of 1,000 samples (a one-second window) in steps of 500 ms. A candidate event was flagged each time LL exceeded 1.5× median for each channel. A total of 27,079 candidate events were identified.

Multitaper spectral estimation using a sliding 200-ms window with 10-ms steps and 5 Hz smoothing was performed to construct spectrograms to visualize µECoG frequency changes over time. Spectrograms were then z-scored to normalize power across each channel. Traces (with and without local re-referencing) and normalized spectrograms were evaluated by a board-certified epileptologist (D.F.) and a trained reviewer (J.S.). Events were labeled as microseizures if they were deemed by both reviewers to have met three criteria^16^: 1) paroxysmal start from baseline, 2) evolution in frequency over time, and 3) return to baseline.

To quantify the evolution of frequency over the course of an event, the maximum frequency was plotted against time. The maximum frequency over a time bin *t* for a power spectral estimate *P*(*f*), where *f* is frequency, is defined as:

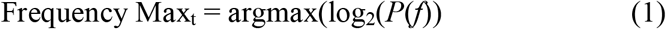

The rate of frequency change for a given event was calculated as the difference in Frequency Max at the start and end of the event divided by the duration.

To investigate the effect of contact size on microseizure detection, adjacent electrodes on the 244-channel array were averaged to simulate larger electrodes. The LL ratio—the LL over a short-term window (the preceding 1 s) divided by a long-term window (the preceding 60 s) calculated in steps of 50 ms—was used to illustrate the sensitivity of microseizure detection at different contact sizes.^13,25^ The microseizures at each contact size were labeled as described above based on spectrograms that were constructed using the averaged traces. To investigate the effect of contact spacing on microseizure detection, electrodes on the 244-channel array were removed to simulate arrays of varying pitch. The microseizures on the channels that remained were then counted.

### Statistical methods

Multiple statistical tests were performed to assess whether the microseizure rate within the EC in awake and anesthetized epilepsy patients differed. A two-sided permutation test (*n*=10,000 permutations) was used to test for a difference in means; a two-sided Mann-Whitney U test was used to test for a difference in ranks; and a two-sided Kolmogorov-Smirnov test was used to test for a difference in distributions. To assess whether the microseizure rate in the recordings obtained within the EC (pooled across awake and anesthetized patients) was higher than the microseizure rate in the recordings obtained in non-EC (in epilepsy and movement disorder patients), one-sided permutation (*n*=10,000), Mann-Whitney U, and Kolmogorov-Smirnov tests were used. A one-sided permutation test (*n*=10,000) was performed to test the null hypothesis that the mean power in the low-frequency (<30 Hz) or high-frequency (70–150 Hz) band for a single channel was higher than the mean power across all channels involved in the event. Significance for all tests was defined as *P*<0.05. For multiple comparisons, the Benjamini-Hochberg procedure for controlling the false discovery rate was used with an alpha value of 0.01. Statistical analyses were performed using MATLAB 2021a (MathWorks, Inc.; Natick, MA).

### Data availability

Data obtained in this study are available upon request. Please contact the corresponding authors with any inquiries.

## Results

We performed intraoperative recordings using μECoG arrays (median duration: 7.6 min, range: 5.0–53.7 min) in nine patients with focal epilepsy and two patients without epilepsy (Table 1). In two epilepsy patients (P5 and P7), we obtained recordings from cortex not involved in initiation or early spread of seizures as defined by prior invasive EEG monitoring. Fig. 2 shows representative recordings from the left posterior superior temporal gyrus of a patient with focal onset epilepsy (P1), obtained using the 256-channel µECoG array. As in previous reports of extra-operative recordings performed in awake patients,^16,22^ we identified microseizures that localized to single electrodes in an anesthetized patient (Fig. 2B-E). Unexpectedly, we found that the microseizures occurred in close spatial and temporal proximity (Fig. 2B,D), suggesting propagation of microseizure activity on a millimeter scale over the course of seconds. We confirmed with time-frequency analysis that the detected microseizures displayed the stereotypical characteristics of an electrographic seizure: 1) paroxysmal start, 2) spectral evolution, and 3) return to baseline (Fig. 2E).

**Figure 2.**
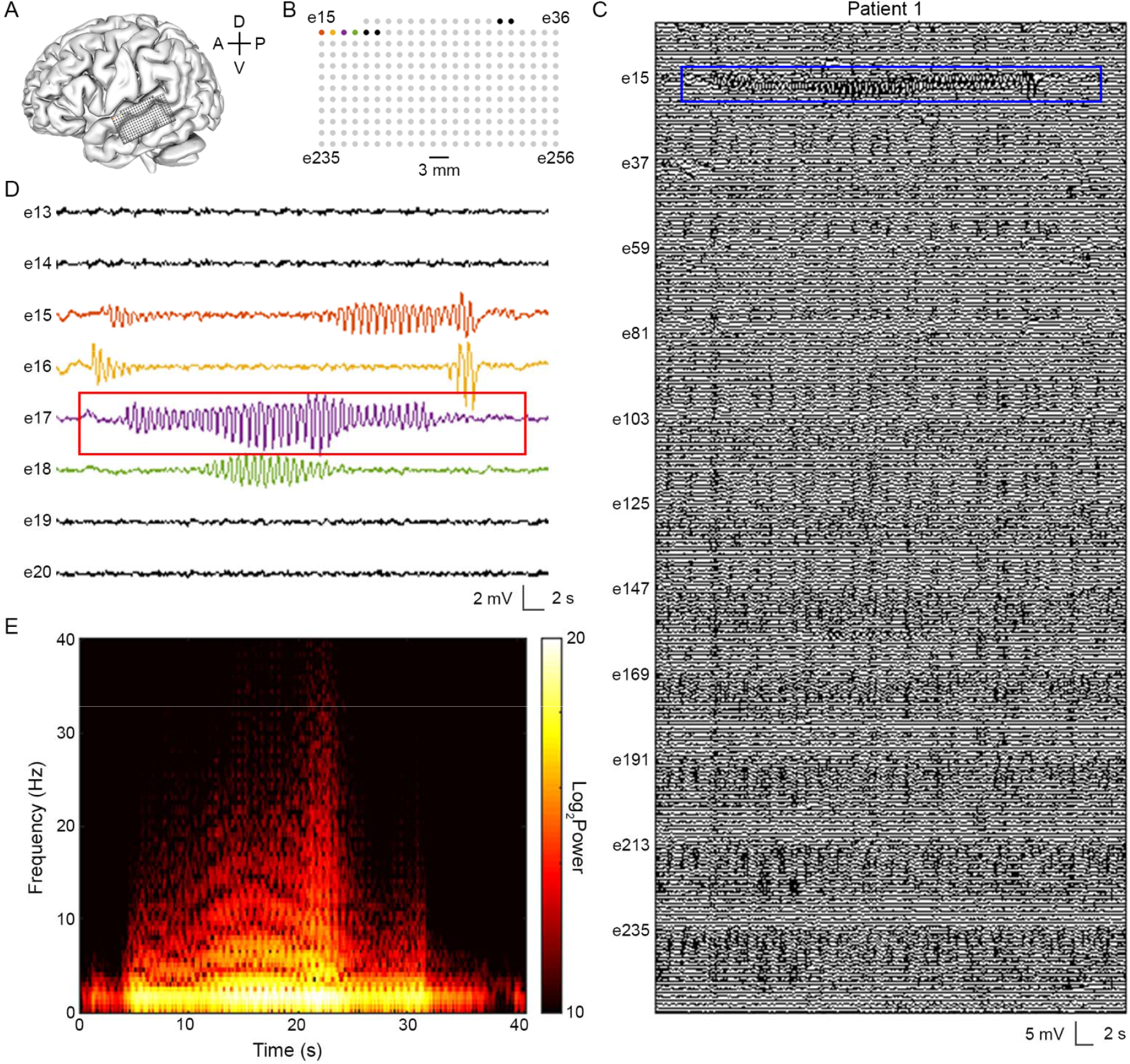
A high-density µECoG array enables detection of electrographic seizure activity limited to single electrodes. A) Schematic of array recording location in patient P1. A: anterior, P: posterior, D: dorsal, V: ventral. B) Map of electrode contacts in 256-channel array. Colored contacts correspond to single channels where microseizure events were detected. C) µECoG signal across all 256 channels. Blue outline delineates zoom window shown in panel D. D) µECoG signal showing electrographic seizure activity in electrodes 15, 16, 17, and 18. Red outline delineates spectrogram window shown in panel E. E) Spectrogram of the microseizure event found on electrode 17. The event demonstrates hallmarks of electrographic seizure activity: paroxysmal change from background, temporal and spectral evolution, and discrete termination.

Of the candidate events identified with the LL detector across 13 recordings in 11 patients, we identified 143 microseizures. Nearly all events (98.6%) occurred in the seven of nine epilepsy patients in which recordings were performed within the clinically-defined EC (Fig. 3A). For the two epilepsy patients (P5 and P7) in which recordings were performed distal to the EC, we did not observe any microseizures. We additionally detected two microseizures in a movement disorder patient (P10). The means, ranks, and distributions of microseizure rates in awake (*n*=3) and anesthetized (*n*=6) epilepsy patients were not significantly different (permutation test, test statistic = 2.0483, *P*=0.5703; Mann-Whitney U test, *U*_*1*_=12.5, *U*_*2*_=5.5, *P*=0.6190; Kolmogorov-Smirnov test, *D*\*=0.5, *P*=0.5344) (Supplementary Fig. 1); therefore, we pooled recordings from awake and anesthetized epilepsy patients into a single group. The microseizure rate from recordings within the EC (*n*=9) was significantly higher than in non-EC controls (*n*=4) in means, ranks, and distributions (2.01 vs. 0.01 microseizures/min, permutation test, test statistic = 2.0028, *P*=0.0068; Mann-Whitney U test, *U*_*1*_=5, *U*_*2*_=31, *P*=0.0210; Kolmogorov-Smirnov test, *D*\*=0.7778, *P*=0.0159) (Fig. 3B). These results suggest that the presence of microseizures may serve as a potential biomarker for EC.

**Figure 3.**
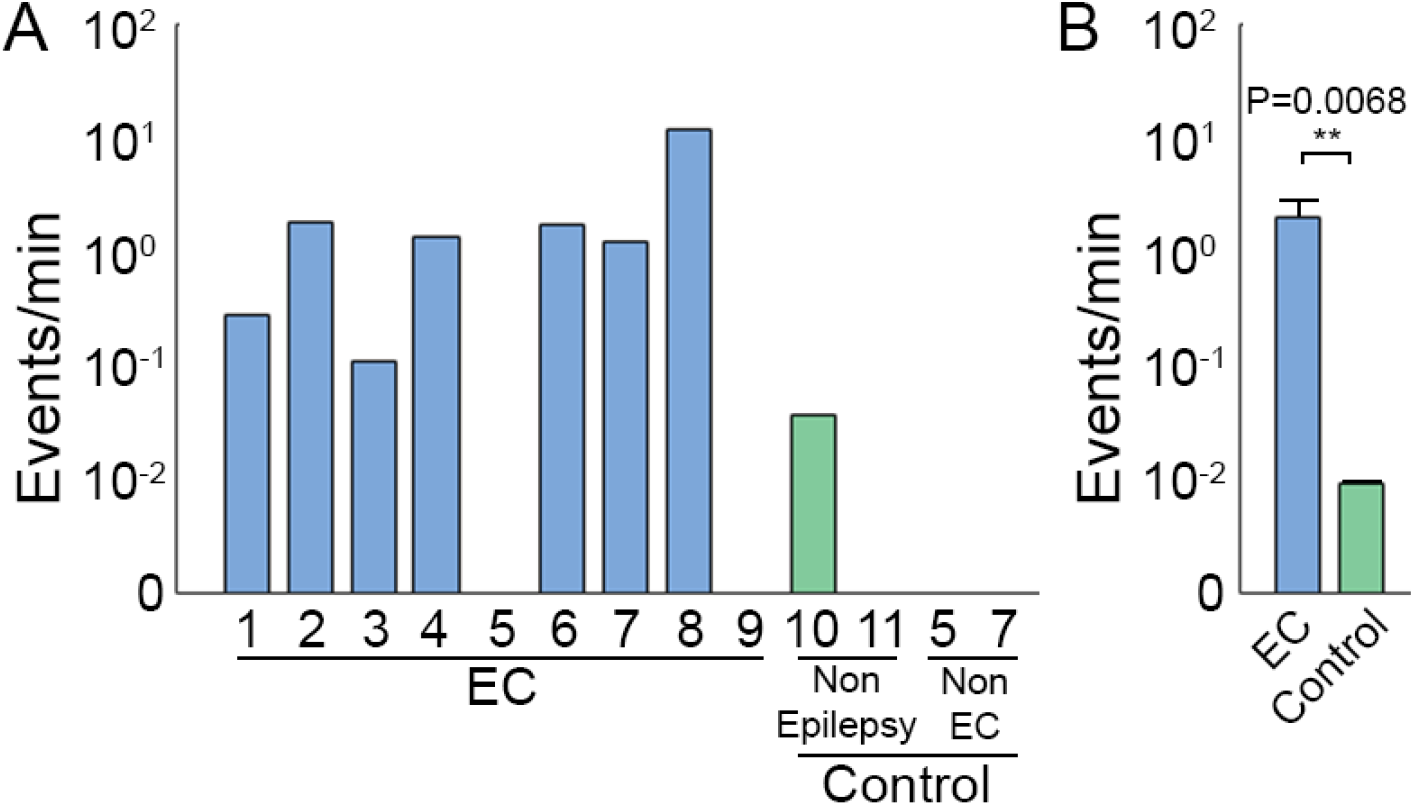
Microseizure rate is higher in clinically identified epileptic cortex than in cortex areas not involved with seizure onset or early spread. A) Microseizure rate observed within epileptic cortex (EC) in epilepsy patients (blue) and within non-EC in movement disorder and epilepsy patients (green). B) Mean microseizure rate observed across all recording sites in EC (*n*=9, blue) and across control sites (*n*=4, green). Permutation test, \*\**P*<0.01.

We identified diverse spatial and temporal patterns of microseizure activity in epilepsy patients (Fig. 4). The mean microseizure duration was 5.91 s (standard deviation: 11.4 s, range: 0.2–106.6 s). The mean number of channels involved per microseizure event was 3.0 (standard deviation: 5.8, range: 1–29). Unlike in patient P1, in which microseizures appeared on nearby electrodes over the course of seconds, the spatially-clustered microseizures detected in patients P4 and P7 appeared within milliseconds across channels (Fig. 4A-J). In patient P8, microseizure events were identified on a single channel without any microseizure activity detected in neighboring channels (Fig. 4K-O). Quantification of frequency evolution over the course of these events revealed distinct patterns (Fig. 4E,J,O). The rates of change of Frequency Max were -15.3, 159.7, and -7.8 Hz/s for the microseizures in patients P7, P4, and P8, respectively.

**Figure 4.**
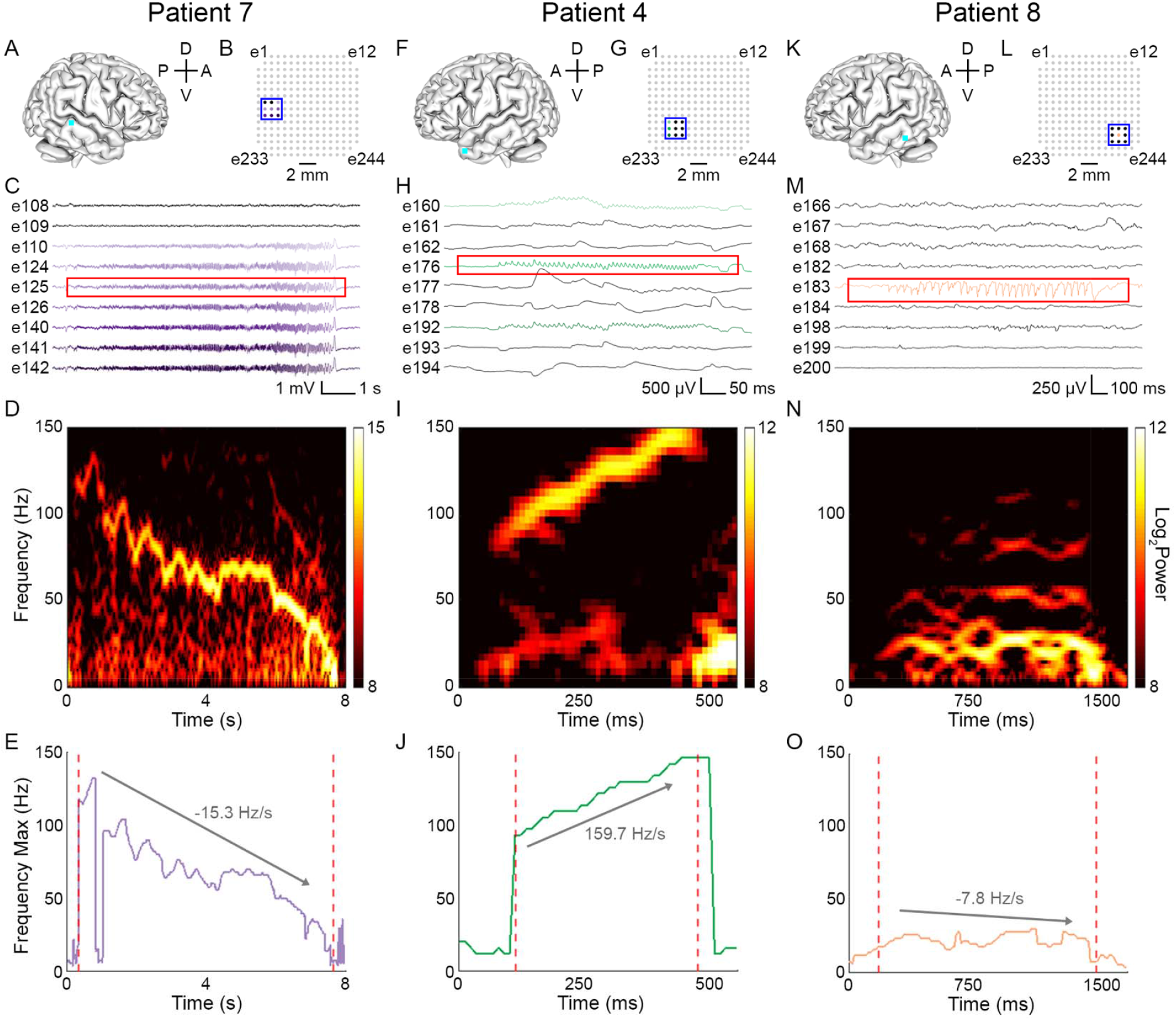
Microseizures in epilepsy patients vary in spatial extent, duration, and frequency. A,E,I) Schematic of array recording location (cyan square) in patients P7 (A), P4 (E), and P8 (I). A: anterior, P: posterior, D: dorsal, V: ventral. B,F,J) Map of electrode contacts in 244-channel array. Colored contacts correspond to single channels where microseizure events were detected. Blue outline delineates the channels whose traces are shown in panels C, G, and K. C,G,K) µECoG signal showing microseizures. Colored traces indicate channels where events were detected. Red outline delineates spectrogram window shown in panels D, H, and L. D,H,L) Spectrogram of the microseizures boxed in red in panels C, G, and K. M,N,O) Frequency Max of microseizures displayed in panels D, H, and L. Dotted red lines indicate start and end of microseizure. Labeled arrow shows change in Frequency Max from start to end of event.

We next sought to determine the extent to which contact size and density contributed to microseizure event detection. We simulated recordings from virtual electrodes with contact diameters of 1.7, 2.4, and 3.2 mm by spatially averaging the signals from adjacent electrodes (Fig. 5A). Figure 5B shows an example isolated microseizure event that was not detected as the virtual electrode contact diameter increases. The microseizure was only detectable when recorded with a single microelectrode. Larger virtual electrodes simulated through spatial averaging were unable to detect this event. Since conventional macroelectrode grids have a contact diameter of 2.3 mm, this example highlights the potential for electrodes with large contact sizes to fail to detect highly focal events, such as microseizures.

**Figure 5.**
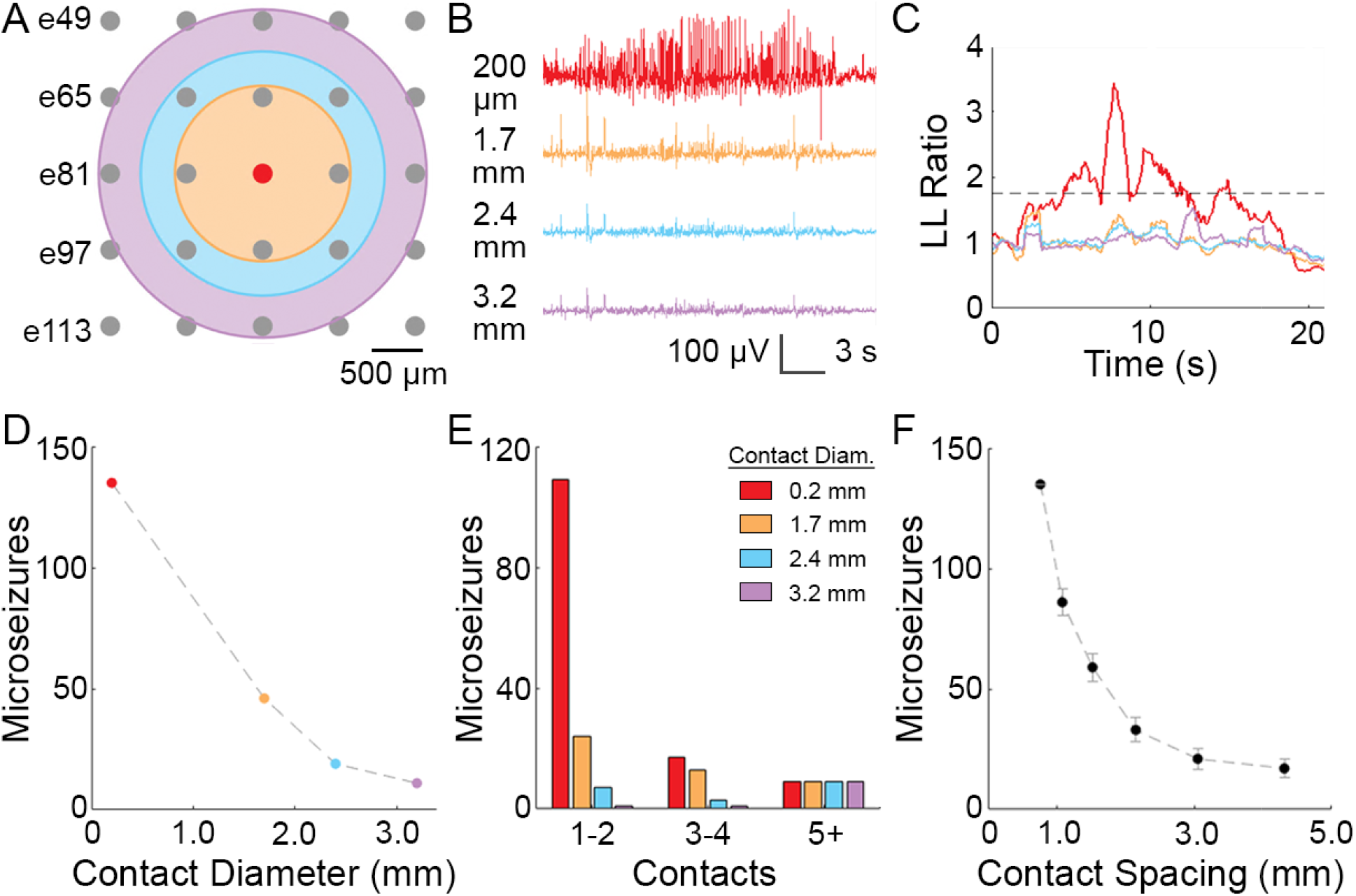
Increased spatial resolution via smaller contacts at greater density facilitates microseizure detection. A) Map of electrode contacts in 244-channel µECoG array. Spatially-averaged electrodes with diameters of 1.7 mm (orange), 2.4 mm (blue), and 3.2 mm (purple) are shown. B) Spatially-averaged µECoG signal shown in colors corresponding to electrodes from panel A. Red trace highlights microseizure event isolated to a single channel. C) Line length (LL) ratio for spatially-averaged traces in panel B. Dotted line shows threshold LL ratio of 1.75. D) Total microseizures detected by 244-channel arrays at various contact diameters. Colors correspond to electrode schematic in panel A. E) Total microseizures detected at various contact diameters, grouped by the number of contacts per microseizure. F) Total microseizures detected by 244-channel arrays at different spacing widths (center-to-center). Error bars represent standard deviation assuming detection of microseizures follows a Bernoulli process.

We analyzed virtual recordings across five epilepsy patients (P2, P4, P6, P7, P8) of all 135 microseizure events acquired using the highest-density 244-channel µECoG array. We found that the number of microseizures detected by virtual electrodes declined with increasing simulated contact diameter: 46 events (1.7 mm contact diameter), 19 (2.4 mm), 11 (3.2 mm), from the set of 135 events (Fig. 5D). Our data suggest that ∼86% of microseizures fail to be detected at 2.4 mm, the contact diameter closest to macroelectrodes used in conventional clinical subdural grids. As expected, events that were detected on 1-2 contacts exhibited the sharpest decrease (93.6%) from 0.2 to 2.4 mm contact diameter (109 to 7 microseizures), while events that were more spatially distributed (i.e., appeared on >5 contacts) were more likely to be captured with electrodes with larger simulated contact sizes (Fig. 5E). We also performed analyses to investigate the effect of contact density on microseizure event detection (Fig. 5F). By removing channels from our analysis, we simulated recordings from arrays with increased contact spacing. Consistent with the presence of a spatially-focal event, the number of detected microseizure events decreased with increasing contact spacing: 86 events (1.08 mm spacing), 59 (1.52 mm), 33 (2.15 mm), 21 (3.05 mm), and 17 (4.31 mm), out of a total of 135 events observed on the full array.

Finally, we asked whether the µECoG array could identify the putative focus of spatially distributed events. Of the 143 microseizure events detected, nine (6.3%) occurred simultaneously on more than five contacts. We show one example in patient P4 that was recorded across eight channels (Fig. 6A-C). We first analyzed the low-frequency power (<30 Hz), a frequency range traditionally used by clinicians to identify electrographic seizure activity (Fig. 6D). Low-frequency local field potential (LFP) power (normalized to a 5 s baseline period) in two electrodes, e176 and e192, was significantly greater than the mean across all eight electrodes (e176: test statistic = 0.1131, *P*<1e-4, e192: test statistic = 0.0353, *P*<1e-4; random permutation test). Multiple sources contribute to low-frequency LFPs; however, the high-frequency extracellular potential is known to reflect locally-generated neural activity.^26^ Therefore, we also examined power in the high-gamma band (70–150 Hz) (Fig. 6E). Electrodes e176 and e192 had significantly elevated high-gamma power compared to the mean across all eight electrodes (e176: test statistic = 0.2010, *P*<1e-4, e192: test statistic = 0.0864, *P*<1e-4; random permutation test). This result further supports the idea that the putative focus of the microseizure event is spatially localized and near the two recording sites (Fig. 6F).

**Figure 6.**
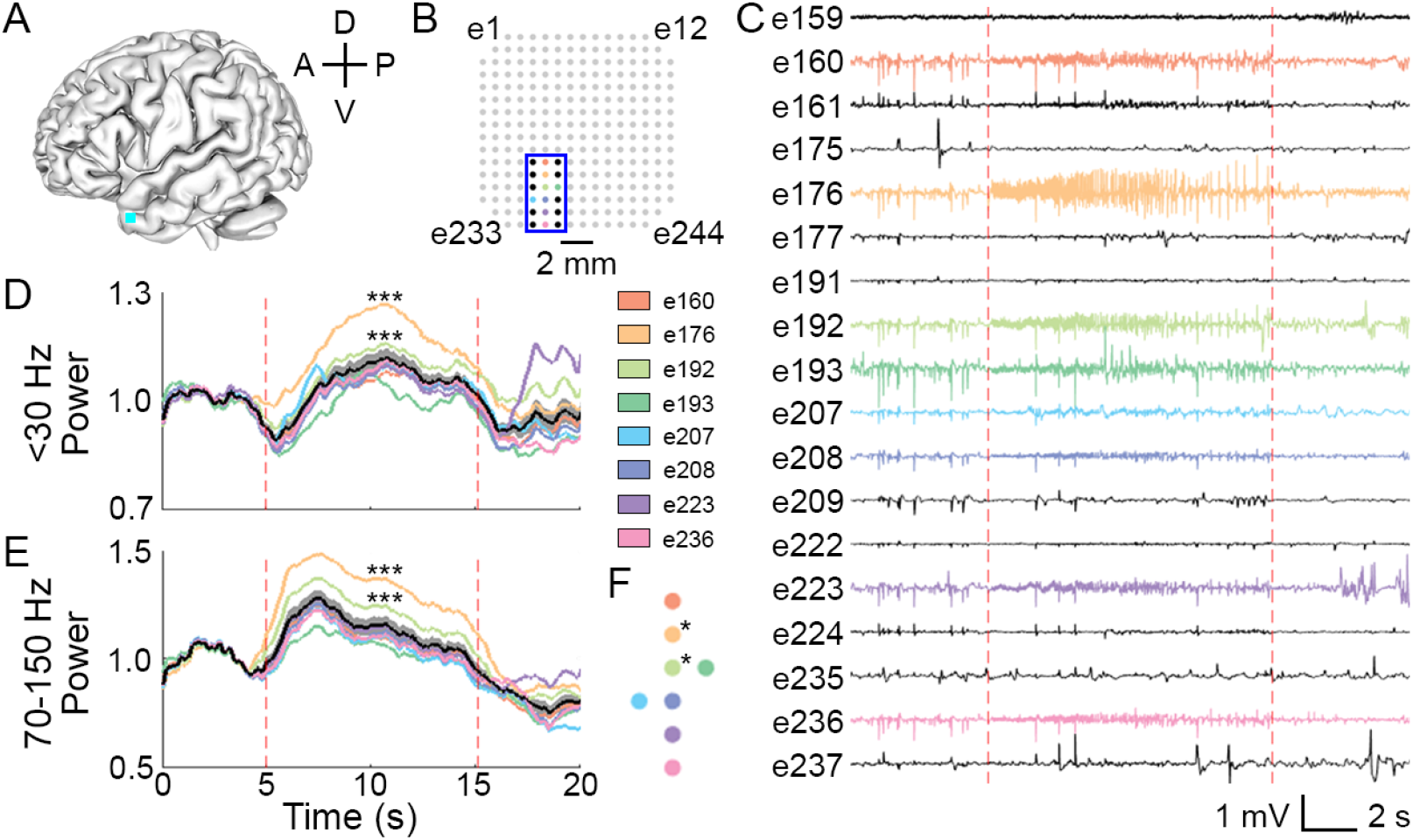
Spectral analysis of spatially distributed events highlights potential to identify the focus of activity within a microseizure. A) Schematic of array recording location (cyan square) in patient P4. A: anterior, P: posterior, D: dorsal, V: ventral. B) Map of electrode contacts in 244-channel array. Colored contacts correspond to single channels where microseizures were detected. Blue outline delineates the channels whose traces are shown in panel C. C) µECoG signal showing microseizures. Colored traces indicate channels where events were detected. Dotted red lines indicate start and end of event. D) Low-frequency (<30 Hz) and E) high-gamma (70–150 Hz) power derived from summed multitaper spectral estimates, normalized to the baseline period of 5 s preceding the start of the microseizure. The mean power (+/- SEM) is shown in black (grey shading). Permutation test, \*\*\**P*<0.0001. F) Map of electrode contacts where microseizures were detected. Asterisks (*) signify electrodes which had significantly elevated power compared to the mean.

## Discussion

Our results show that microseizures are a commonly occurring phenomenon in epileptic patients, are specific to epileptic cortex (EC – an area defined by clinical seizure onset or early spread) and can be detected with high-density µECoG arrays during brief interoperative recordings. We found that high-density µECoG arrays with broad spatial coverage can detect microseizures that occur in the absence of clinical seizures in both anesthetized and awake epilepsy patients. Detected microseizures had variable frequency dynamics, duration, and spatial extent, and occurred more frequently in epilepsy patients than in controls (*P*=0.0068; Fig. 3). Our results also suggest that standard clinical arrays would not have detected most microseizure events (Fig. 5). Specifically, spatial averaging and subsampling simulations indicated that increases in contact size and spacing would result in fewer detectable microseizure events across all recordings. This suggests that µECoG arrays with small contacts, high contact density, and large coverage are crucial to capturing microseizure activity. We also demonstrated the utility of µECoG arrays to localize possible foci of microseizures occurring across an area of multiple channels based on differences in low frequency (<30 Hz) and high-gamma (70–150 Hz) signal power between contacts (Fig. 6). Our findings show that microseizures are specific to EC but would be undetectable by clinical macrocontacts, and that high-density µECoG arrays with broad spatial coverage are essential to their detection.

Our µECoG arrays offer several strengths that enabled the capture and spatiotemporal analysis of microseizures in the human brain. A crucial advantage is the balance between spatial resolution (contact size and density) and spatial coverage. Most prior research on seizure activity in human subjects has used data collected from either clinical grids with coarse spatial resolution (10-mm pitch) but extensive spatial coverage (on the order of 6,400 mm^2^), or very high-resolution microelectrode arrays with limited coverage, such as Utah arrays with penetrating silicon shanks (0.4-mm pitch, 16-mm^2^ coverage) or the cut tips of microwires (1-mm pitch, ≤10 clusters of ≤16-mm^2^ coverage each) protruding between standard clinical grid or sEEG electrode contacts.^16,22^ Here we describe devices and methods that yield microscale recordings while maintaining extensive spatial coverage comparable to clinical grid arrays. Although the contact size and spacing of our µECoG devices do not allow for the study of microseizures in relation to spiking activity as captured by penetrating microelectrode arrays, our spatial coverage and standard ECoG signal analysis methods may enable more direct and intuitive clinical translation of our results. Another key advantage of using these arrays is the choice of the LCP-TF material and electroplated Au and PtIr microcontacts. Our flexible LCP-TF electrode arrays better conform to the curvature of the cortex than penetrating microelectrode arrays and have smoother surfaces, offering higher signal-to-noise and increased protection of brain tissue. Previous work using penetrating arrays or cut wire tips has received some scrutiny due to the similarity between reported microseizures and electrographic signals resulting from cortical injury.^27,28^ The smooth surface of our µECoG arrays mitigates this concern, providing greater confidence that the microseizures captured in our recordings are truly electrographic events and not a result of tissue injury.^23^ These qualities of our LCP-TF µECoG arrays, as well as the small contact size, high contact density, and broad coverage, are crucial for recording microseizures.

Interestingly, the occurrence of microseizures in microarray recordings has varied between studies and devices. Using penetrating platinum-coated silicon microelectrodes with 3–5 µm tips spaced 400 µm apart, Schevon et al.^22^ found microseizures in three of five epileptic patients (60%). Using an array of 30-µm diameter PEDOT:PSS contacts spaced 50–600 µm apart, Yang et al.^19^ observed microseizure activity in only 1 of 30 epilepsy patients (3%). In contrast, Stead et al.^16^ used the cut ends of PtIr wire as electrode contacts with 40-µm diameter with 0.5–1 mm spacing and observed microseizures in 14 of 14 epilepsy patients (100%). Although we used devices with larger contact diameter (200 µm) and larger spacing (0.762–1.72 mm) than in these previous studies, our devices also provided the largest coverage (144–798 mm^2^). We observed microseizures in 7 of 9 epilepsy patients (78%), and these events were detected at an average rate of 2.01 events/min (Fig. 3). The event frequency we observed is greater than previously reported in a similar study^16^ (0.014 microseizures/min). This variability in microseizure frequency between studies may be due to differences in the brain regions recorded, patient disease states, microelectrode array coverage and pitch, microseizure determination during visual review, anesthetic regimens, and states of anesthesia and wakefulness. For example, some anesthetics such as remifentanil and sevoflurane may promote interictal epileptiform activity while others such as isoflurane may suppress interictal epileptic activity.^29^ We recorded from patients under various anesthetics including remifentanil, sevoflurane, and isoflurane (Table 1). However, we did not find a significant difference between the frequency of microseizures in epileptic patients under anesthesia at the time of recording and epileptic patients who were awake (Supplementary Fig. 1). Further work recording at high resolution across a variety of epileptic pathologies is needed to better understand the variability in microseizure occurrence between patients and to better identify the smallest region within which clinically relevant microseizure events occur.

A key result of our analysis is that clinical macro contacts would be unlikely to detect the majority of microseizure events recorded by our LCP-TF µECoG arrays. LFP signals captured by metal contacts placed on the brain surface predominantly reflect a summation of postsynaptic currents in mostly superficial cortical layers.^20,26^ Larger contacts measure an average of activity across a greater number of neuronal sources. We simulated the signal that larger contacts would measure during microseizures by averaging signals captured between multiple microcontacts in our recordings. While some features of the signal are visible across spatial scales, the microseizure quickly becomes undetectable when larger contacts are simulated. When simulating the signal captured over a 2.4-mm diameter area, a size comparable to clinical standard 2.3-mm diameter contacts, we detected 86% fewer microseizures than when using 200-µm diameter contacts. Although spatial averaging between microcontacts is a reasonable approximation, it is not necessarily equivalent to the signal that would be measured by a larger solid contact. Specifically, our simulated result does not account for the LFP signal between the microcontacts or the decrease in impedance and change in signal-to-noise ratio when using a contact with greater surface area. However, assuming that the electric field in the <300 Hz frequency range is smooth between recording sites, our spatial averaging analysis provides a useful proxy for the signal observed when using larger area contacts.^30^ This result is notable given that the majority (81%) of microseizures detected in our recordings occurred on only one or two contacts at a time.

We also found that the density of microcontacts was critical to detecting microseizure events (Fig. 5). By sub-sampling the array, we were able to simulate recordings of varying contact spacing and observed that fewer microseizures were detected as contact density decreased. More notably, the decline in the number of microseizure events detected (from 135 to 86 events) between 0.76-mm and 1.08-mm contact spacing suggests that an even greater density of contacts (<0.76-mm spacing) could result in a substantial increase in the number of detected microseizure events. These results support our hypothesis that µECoG arrays capture epileptic events that would not be observed using standard clinical grids. Determining the smallest spatial scale needed to capture all microseizure events requires further microscale study of epileptic cortex, along with technology development and biophysical modeling of µECoG recordings.^31,32^ As thin film fabrication and encapsulation of actively-multiplexed microelectrode arrays continue to improve, spatial resolution and coverage of recording arrays will increase, enabling more comprehensive recordings of microscale epileptic activity.^33^

Identifying foci of activity within larger seizure events is essential for effective therapeutic targeting. While most microseizure events occurred on only 1-2 channels, we also identified events that were spatially distributed over neighboring channels (Fig 6). In such cases, we investigated whether there was an identifiable focus of activity. Traditionally, epileptologists have assessed low frequency features (<30 Hz) of ECoG recordings to identify seizure activity. However, recent work has demonstrated that high-gamma LFP signals (70–150 Hz) reflect more spatially localized spiking activity.^34,35^ In the case of spatially distributed microseizures in our recordings, particular channels had signal power within both these frequency ranges that was significantly elevated compared to the mean across neighboring channels involved in the microseizure. We hypothesize that the channels with higher signal power, especially in the high-gamma frequency range, are distinctly active within the distributed microseizure events and thus could be a more optimal therapeutic target, especially in the case of micro-stimulation.

Some limitations of the current study can be addressed by future work. For example, we gathered only brief intraoperative recordings, often while the patient was anesthetized (Table 1). This experimental setup limits our ability to relate these intraoperative microseizures with macroscale seizures captured during awake clinical monitoring. However, the ability to capture microseizure activity from intraoperative recordings also presents a promising biomarker for epileptogenic cortex which may improve intraoperative delineation of EC. With the opportunity to record during semi-chronic implantation, Schevon et al.^22^ and Stead et al.^16^ observed the evolution of microseizures into clinical seizures, thus relating the two phenomena in a meaningful way. Further data collection, particularly recording simultaneously at both the micro- and macroscale with wide coverage during short-term implantation, will be needed to identify the smallest spatial scale at which seizure activity initiates and propagates.^23^ Data from hybrid-scale recordings would answer questions regarding the relationship between micro- and macroscale seizures and how they may best be interrupted by treatments including resective surgery, laser ablation, and RNS. In addition, we have collected microscale recordings only from the surface of the brain; it is not yet known to what extent µECoG recordings reflect activity from deeper neuronal sources. The source of microseizures and their occurrence and propagation among various cortical layers remains unclear. Future work in which laminar depth recordings are conducted simultaneously with microscale surface recordings will be critical to understanding the relationship between microseizures captured at the brain surface and activity from deeper structures.^24^ While grids remain a crucial tool for clinical delineation of EC in presurgical and intraoperative evaluation, there is expanding use of sEEG electrodes in clinical monitoring due to their improved safety and comfort profiles.^21^ Therefore, micro-sEEG devices are also needed to provide additional clinically relevant information on microseizures in epilepsy.^23^

This study and others raise the question as to why microseizures are found not only in epileptic patients but also occasionally in control patients.^16^ There have been reported cases of cryptogenic epilepsy in Parkinson’s patients, so the presence of microseizures in movement disorder patients may reflect an underlying risk for epilepsy.^36^ It may also be that microseizures occur spontaneously in nonepileptic brain tissue, but that differences in connectivity or inhibition between neural populations of healthy versus pathologic tissue influence the rate of occurrence and propensity of microseizures to spread into large-scale seizures.

Our results demonstrate that microseizures are specific to EC, and that high-density microcontact arrays with extensive coverage are crucial to capturing microseizures, as most events would be undetectable on clinical macro contacts. We have successfully identified microseizure events in our intraoperative recordings of epileptic and non-epileptic patients using flexible LCP-TF µECoG arrays. We found elevated rates of microseizure events in epileptic cortex and demonstrated the utility of dense, microscale recording arrays to capture these events. Our results contribute to a growing body of evidence that epileptic activity in the human brain is occurring at much finer spatial scales than reflected in clinical-standard ECoG and sEEG recordings. Furthermore, we show variations in high-gamma power between neighboring contacts during microseizures which differentiate even more specific foci within microseizure events. Since the success of surgical treatments such as resection, laser ablation, and RNS are dependent on accurate targeting of seizure initiation sites, precise localization of EC is critical to improving outcomes for patients with drug-resistant epilepsy. This work demonstrates the importance of additional research on microscale phenomena in epilepsy, particularly using µECoG arrays with both high resolution and broad coverage. We expect that further research will reveal how microscale biomarkers of EC may be used for differentiating epileptic from non-epileptic tissue in cases of ambiguity, thus improving therapeutic outcomes in refractory epilepsy.

## Abbreviations

DBS: deep brain stimulation;
EC: epileptic cortex;
ECoG: electrocorticography;
EZ: epileptogenic zone;
IRB: Institutional Review Board;
ISO: International Organization of Standardization;
LCP: liquid crystal polymer;
LCP-TF: liquid crystal polymer thin-film;
LFP: local field potential;
LL: line length;
PEDOT:PSS: poly(3,4-ethylenedioxythiophene) polystyrene sulfonate;
PtIr: platinum-iridium;
RNS: responsive neurostimulation;
sEEG: stereo-electroencephalography;
TF: thin film;
VNS: vagal nerve stimulation;
µECoG: micro-electrocorticography;
µHDMI: micro high-definition multimedia interface

## Acknowledgements

We would like to thank the patients who participated in this study, as well as Lora Fanda and Beenish Mahmood, who assisted with data collection at NYU Grossman School of Medicine, and Anna Thirakul, who assisted with data collection at Duke University School of Medicine. We would also like to thank Finding a Cure for Epilepsy and Seizures (FACES) for providing financial support.

## Funding

J.S. receives support from the NIH (T32 GM136573), the Vilcek Fellowship, and the American Epilepsy Society. D.F. receives support from the NIH (R01 NS06209207, NIH R01 NS109367, CDC U48DP006396-01SIP 19-003) and Epitel. O.D. receives support from the NIH (R01 MH107396, U01 NS099705, R01 MH111417, U01 NS090415, R01 MH116978, R01 HL151490), Tuberous Sclerosis Alliance, Epilepsy Foundation of American, and the National Science Foundation. W.D. receives support from the NIH (R01 NS062092, R01 MH116978). S.D. receives support from the NIH (RF1 MH116978, R01 MH111417, R01 NS062092), Office of Naval Research, and the Templeton World Charity Foundation. BP receives support from NIH U01 NS099697, U01 NS099577, U01 NS103518, U01 NS122123, R01 NS104923, ARO 68984-CS-MUR, NSF IOS-1557886 and NSF IIS-2113271. J.V. receives support from Grant #DoD EP200077, NIH U01 NS099697, NIH 1UG3NS120172, NIH 1U01NS123668, NSF CBET-1752274, and NIH CTSA grant UL1TR002553. D.S. receives support from the NIH (K12 NS080223) and the Esther A. & Joseph Klingenstein Fund.

## Competing interests

Parts of the technology described here are patent pending under ‘Electroencephalography (EEG) Electrode Arrays and Related Methods of Use’ U.S. Patent Application # PCT/US2020/051400. F. Solzbacher declares financial interest in Blackrock Microsystems LLC and Sentiomed, Inc., managed by University of Utah’s COI management. W. Doyle and D. Friedman declare financial interest in Neuroview Technology overseen by NYU Grossman School of Medicine’s COI management. D. Friedman also receives salary support for consulting and clinical trial related activities performed on behalf of The Epilepsy Study Consortium, a non-profit organization. Dr. Friedman receives no personal income for these activities. NYU receives a fixed amount from the Epilepsy Study Consortium towards Dr. Friedman’s salary. Within the past two years, The Epilepsy Study Consortium received payments for research services performed by Dr. Friedman from Alterity, Baergic, Biogen, BioXcell, Cerevel, Cerebral, Jannsen, Lundbeck, Neurocrine, SK Life Science, and Xenon. Dr. Friedman has also served as a paid consultant for Neurelis Pharmaceuticals and Receptor Life Sciences. OD has equity and/or compensation from Privateer Holdings, Tilray, Receptor Life Sciences, Qstate Biosciences, Tevard, Empatica, Engage, Egg Rock/Papa & Barkley, Rettco, SilverSpike, and California Cannabis Enterprises. He has received consulting fees from GW Pharma, Cavion, and Zogenix. Saurabh R. Sinha has received salary/research support for clinical trials from Eisai, Monteris, Neuropace, UCB, and Sunovion. Within the past two years, Dr. Sinha has received payments for consulting/advisory boards from Acquestive, Basilea, Blackthorn Therapeutics, LivaNova, Monteris, Neuropace, SK Lifesciences, and UCB.

## Figures

**Supplementary Figure 1.**
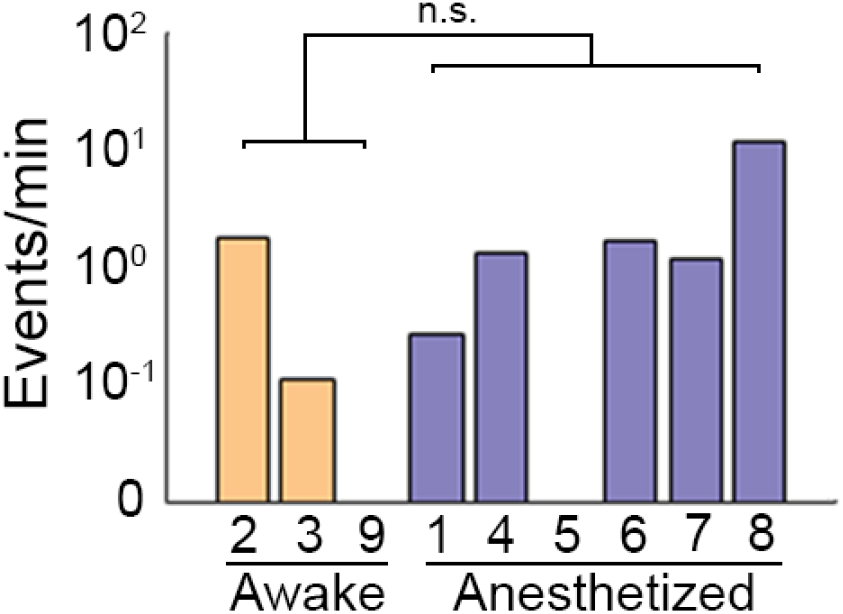
Microseizure rates do not differ significantly between awake (*n*=3) and anesthetized (*n*=6) epilepsy patients. Microseizure rates in awake (P2, P3, P9) and anesthetized (P1, P4, P5, P6, P7, P8) epilepsy patients were compared. Rates did not differ in mean (permutation test, test statistic = 2.0483, *P*=0.5703), rank (Mann-Whitney U test, *U*_*1*_=12.5, *U*_*2*_=5.5, *P*=0.6190), or distribution (Kolmogorov-Smirnov test, *D*\*=0.5, *P*=0.5344). n.s. = not significant.

## Notes

### Author Declarations

Informed consent was obtained from all patients in accordance with the Institutional Review Boards at New York University Langone Health and the Duke University Health System.

